# Plasma p-tau212 as a biomarker of sporadic and Down Syndrome Alzheimer’s disease

**DOI:** 10.1101/2024.10.31.24316469

**Authors:** Przemysław R. Kac, Daniel Alcolea, Laia Montoliu-Gaya, Susana Fernández, Juan Lantero Rodriguez, Lucía Maure, Fernando González-Ortiz, Bessy Benejam, Michael Turton, Isabel Barroeta, Peter Harrison, Laura Videla, Nicholas J. Ashton, Alberto Lleó, Henrik Zetterberg, María Carmona-Iragui, Thomas K. Karikari, Juan Fortea, Kaj Blennow

**Affiliations:** Institute of Neuroscience and Physiology, Department of Psychiatry and Neurochemistry, The Sahlgrenska Academy at University of Gothenburg, Mölndal, Sweden; Sant Pau Memory Unit, Hospital de la Santa Creu i Sant Pau, Biomedical Research Institute Sant Pau, Universitat Autònoma de Barcelona, Barcelona, Spain; Center for Biomedical Investigation Network for Neurodegenerative Diseases (CIBERNED), Madrid, Spain; Clinical Neurochemistry Laboratory, Sahlgrenska University Hospital, Mölndal, Sweden; Bioventix Plc, Farnham, GU9 7SX, United Kingdom; Centre for Age-Related Medicine, Stavanger University Hospital, Stavanger, Norway; NIHR Biomedical Research Centre for Mental Health & Biomedical Research Unit for Dementia at South London & Maudsley NHS Foundation, London, United Kingdom; Department of Old Age Psychiatry, Institute of Psychiatry, Psychology, and Neuroscience, King’s College London, London, London, United Kingdom; Wisconsin Alzheimer’s Disease Research Center, University of Wisconsin School of Medicine and Public Health, University of Wisconsin-Madison, Madison, WI, USA; Hong Kong Center for Neurodegenerative Diseases, Clear Water Bay, Hong Kong, China; Department of Neurodegenerative Disease, UCL Queen Square Institute of Neurology, University College London, London, United Kingdom; UK Dementia Research Institute at UCL, London, United Kingdom; Department of Psychiatry, School of Medicine, University of Pittsburgh, Pittsburgh, PA, USA; Barcelona Down Medical Center, Fundació Catalana Síndrome de Down, Barcelona, Spain; Paris Brain Institute, ICM, Pitié-Salpêtrière Hospital, Sorbonne University, Paris, France; Neurodegenerative Disorder Research Center, Division of Life Sciences and Medicine, and Department of Neurology, Institute on Aging and Brain Disorders, University of Science and Technology of China and First Affiliated Hospital of USTC, Hefei, P.R. China

**Keywords:** Alzheimeŕs disease, Down Syndrome, plasma biomarkers, p-tau212, DYRK1A, DABNI, SPIN, Simoa, CSF biomarkers

## Abstract

**Background:** All individuals with Down Syndrome (DS) will develop full-blown Alzheimeŕs disease (AD) pathology by age 40, decades before the occurrence of sporadic late-onset AD. Understanding this strong biological relation between age and AD pathology risk in DS is important to accelerate diagnostics, disease monitoring, and treatment. Several genes encoded in chromosome 21 including dual-specificity tyrosine phosphorylation-regulated kinase 1A (DYRK1A) have been proven to contribute to the pathology. A recently validated plasma immunoassay to measure tau phosphorylation at threonine-212 (p-tau212) has very high diagnostic accuracy in detecting AD. P-tau212 is also very sensitive to DYRK1A phosphorylation and is increased in DSAD brain lysates. Here, we assessed the potential of this biomarker in DSAD and sporadic AD.

**Methods:** Using Simoa technology, we tested p-tau212 and p-tau181 (n=245 for plasma, n=114 matching cerebrospinal fluid (CSF) samples). We used AUC-ROC to examine diagnostic performance and the DeLong test to compare the AUC-ROC differences between methods. Spearman correlation is used to examine correlations. Fold changes relative to median levels were calculated for their respective asymptomatic groups. ANCOVA followed by Tukey post-hoc test was used to calculate differences across groups. LOESS was used to determine the temporality of plasma biomarker changes.

**Results:** We have confirmed that p-tau212 has extremely high accuracy in detecting AD-related changes in euploid controls. For the DS population, we observed a strong correlation between plasma and CSF p-tau212 (r=0.867; p<0.001). In prodromal DS (pDS) and dementia DS (dDS), we observed significantly elevated levels of p-tau212 in reference to asymptomatic DS (aDS). The diagnostic accuracy to differentiate between aDS and dDS was AUC=0.91 and AUC = 0.86 in discriminating between DS amyloid positive and amyloid negative participants. Plasma p-tau212 started increasing approximately when people became amyloid PET-positive.

**Conclusions:** We have confirmed that the levels of plasma p-tau212 are increased in the DS population and sporadic AD cases including prodromal and MCI states. Plasma p-tau212 might have utility for theragnostic, monitoring therapy efficacy, and as a target engagement biomarker in clinical trials both in sporadic and DSAD.

## Introduction

A triplication of chromosome 21 causes Down syndrome (DS)(1). All individuals with DS develop full-blown Alzheimer’s Disease (AD) pathology by age 40(2) and the lifetime risk of developing AD exceeds 90% in the seventh decade(3). The estimated age of onset of AD dementia in this population is 53.8 years, decades before the occurrence of late-onset AD in the general population(4). Understanding this strong DSAD relationship is important to accelerate diagnostics and treatment. This ultra-high risk is mainly due to the triplication of the amyloid precursor protein (*APP*), which leads to the overproduction of amyloid-β (Aβ) peptides, and increased Aβ plaques formation(5). Although other genes encoded in chromosome 21 such as dual-specificity tyrosine phosphorylation-regulated kinase 1A (*DYRK1A*) and *RCAN1* contribute to the pathology(6–9). *DYRK1A* is a dose-sensitive gene that overexpression contributes to DS cognitive dysfunction(10). This protein phosphorylates different targets involved in AD development and progression for instance Glycogen synthase kinase-3β (GSK-3β)(11), Presenilin1(12), APP(6), and tau(13).

Diagnosing DSAD is challenging due to the association of DS with cognitive dysfunction(14,15). Recent advances allow us to recognize ante-mortem AD pathology using blood-based biomarkers(16–19). These reflect the disease almost as accurately as more expensive and less available CSF and imaging biomarkers(20). Several plasma p-tau biomarkers have been found to reflect both Aβ and tau pathology(21). Recently validated plasma p-tau212 has shown very high performance for detecting those pathologies and AD diagnosis(22). That phosphorylation site might also have the strongest biological correlation with AD pathology in DS since threonine-212 is a primary target for DYRK1A in tau protein(23). Intensified phosphorylation at thr-212 induces tau aggregation, reduces tau binding to microtubules, and increases cell toxicity in in-vitro studies(24). Levels of p-tau212 are highly elevated in reference to AD and control participants in human AD-DS brains(25). Additionally, a major tau phosphatase – Protein phosphatase 1A (PP1A) is not dephosphorylating p-tau212 derived from AD brains(26), suggesting that this epitope might be vulnerable to very subtle changes related to AD pathology. Knowing that direct biological association, we hypothesized that p-tau212 is an accurate tau species to reflect AD pathology in the DS population. To test this hypothesis, we used Single molecule array (Simoa) immunoassays to measure plasma and cerebrospinal fluid (CSF) p-tau212 concentrations in asymptomatic DS (aDS), prodromal DS (pDS), and dementia DS (dDS) individuals, as well as in sporadic AD patients both in Mild Cognitive impairment (MCI) and dementia states and we compare the results with a validated biomarker.

## Methods

### Study design and participants

We performed a cross-sectional cohort study of adults with DS, and euploid individuals along the Alzheimer’s disease continuum in the Hospital of Sant Pau, Barcelona (Spain), Adults with DS in Barcelona were recruited from a population-based health plan designed to screen for AD dementia, which includes yearly neurological and neuropsychological assessments. Those subjects interested in research studies are included in the Down Alzheimer Barcelona Neuroimaging Initiative (DABNI) cohort(14,15). We also recruited euploid controls and sporadic Alzheimer’s disease patients from the Sant Pau Initiative on Neurodegeneration (SPIN cohort)(27).

### Biomarker measurements

Amyloid-β peptides (Aβ40, Aβ42) analyses were performed on the LUMIPULSE G600II, and the cut-off for positivity was defined as in the previous publication(28). All other plasma and CSF biomarker assays were performed on the Simoa HD-X platform at the University of Gothenburg. P-tau212 and p-tau181 concentrations were measured using published and validated in-house assays(16,22). For p-tau181, the AT270 antibody (Invitrogen) was used as a capture antibody and was paired with n-terminal antibody for detection (Tau12; BioLegend). For p-tau212, a sheep monoclonal antibody was used for capture paired with Tau12 as the detector. Coefficients of variation for 3 different internal quality controls for plasma were 5.3-12% for within-plate variation and 6.4%-12% for between-plate variation. For CSF, these values were 10.8%-13.3% and 12.9%-15.5%, respectively.

### Statistical Analysis

We used AUC-ROC to examine diagnostic performance and the DeLong test to compare the AUC-ROC differences between methods. Spearman correlation is used to examine correlations. Fold changes relative to median levels are calculated for their respective asymptomatic groups. Age-adjusted analysis of covariance (ANCOVA) followed by Tukey post-hoc test is used to calculate differences across groups. A First-degree locally estimated scatterplot smoothing curve (LOESS) is independently used in controls and adults with Down Syndrome to determine the temporality of plasma biomarker changes. For Down participants we set the mean age of symptoms onset at 53.8 years according to a previous study from our group(29). All significance tests were two-sided, and significance was set at p<0.05.

## Results

### Cohort characteristics

We tested n=245 plasma samples for p-tau212 and p-tau181. A subset of participants had amyloid PET data. Table 1. shows the demographics, cognitive, and plasma biomarkers across groups of all participants included in the analyses. N=114 (47%) participants had CSF biomarker measurements. Demographics for the CSF subset are shown in Supplementary Table 1.

**Table 1.**
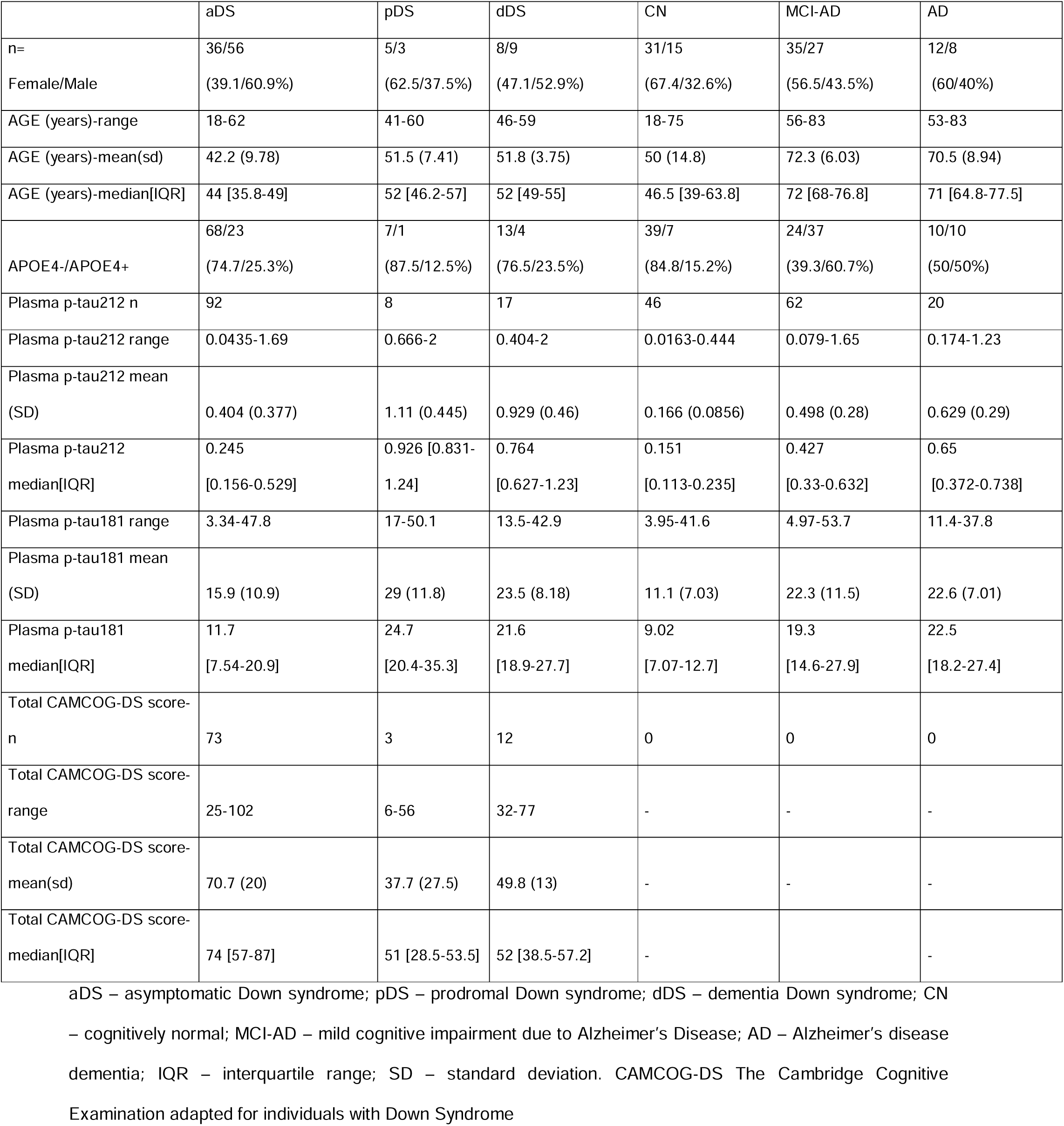
Study Participants.

### Correlations with biomarkers and cognition

Plasma and CSF p-tau212 were highly correlated with each other within the cohort. We observed moderate correlation across sample types in all participants (r=0.712 p<0.0001) (Fig. 1A). However, the strongest correlation of plasma p-tau212 with CSF p-tau212 was observed in the DS-only subgroup (r=0.867, p<0.0001) (Fig 1A). This improved CSF-plasma correlation in DS groups was not seen for p-tau181, which showed similar correlations in the whole cohort and subgroups (r=0.652-0.681, p<0.0001) (Fig 1B). Those results indicate that plasma p-tau212 measurements accurately reflect AD-related p-tau level changes in CSF in sporadic and DS groups.

**Figure 1.**
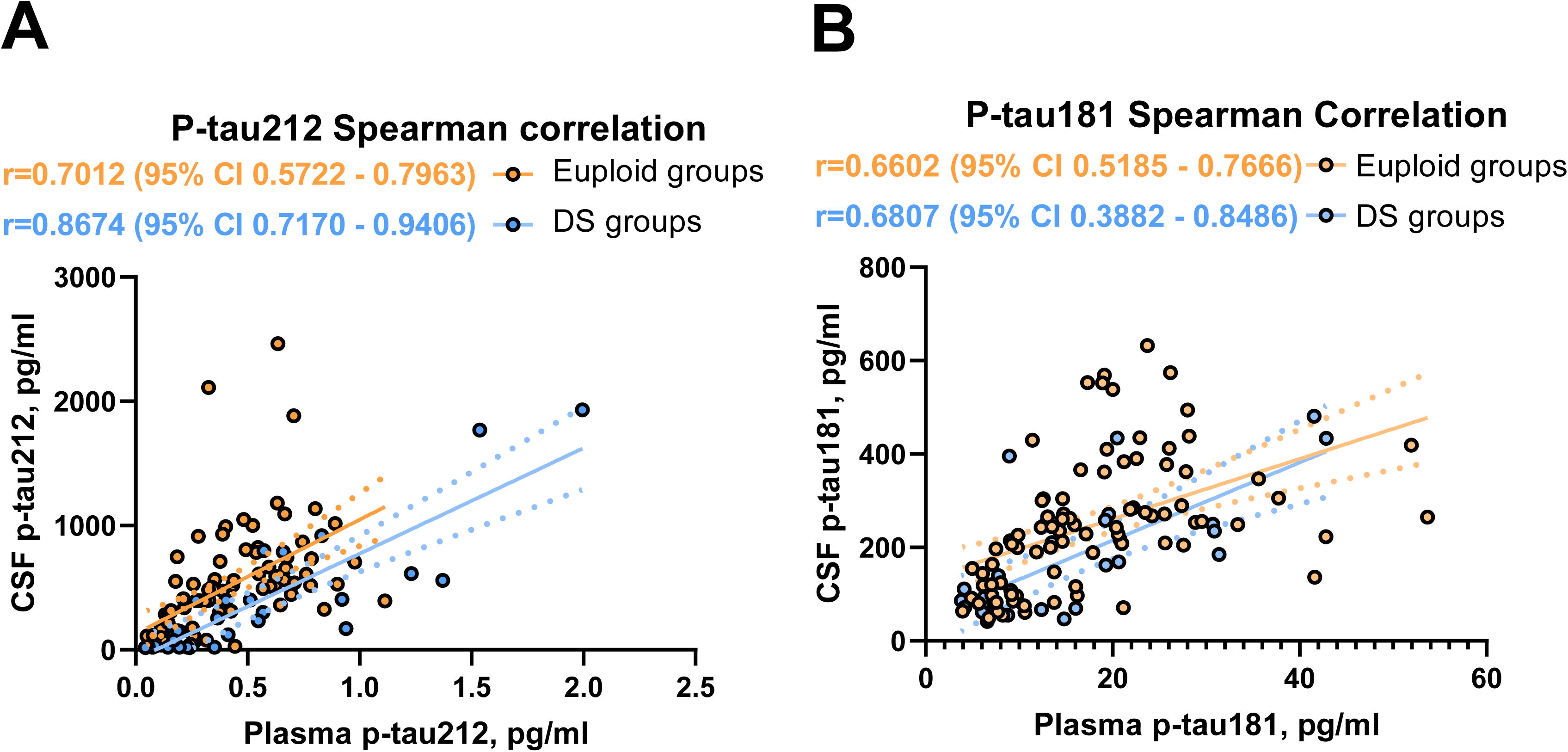
Spearman correlations for plasma and CSF p-tau212 and p-tau181. The Figure shows correlations between plasma and CSF for **A)** p-tau212 and **B)** p-tau181 for euploid groups (n=88; p<0.001; r= 0.701 and r=0.660 respectively) and DS groups (n=26, p<0.001; r=0.867, r=0,681 respectively). The correlation between plasma and CSF measurements for all measurements (n=114, p<0.001) is 0.712 for p-tau212 and 0.652 for p-tau181. The fitted simple linear regression line is presented as a mean and error.

Both biomarkers were correlated with decrease in The Cambridge Cognitive Examination adapted for individuals with Down Syndrome (CAMCOG-DS) in plasma (r=-0.338; p=0.001 for p-tau212 and r=-0.328 for p-tau181; p=0.002;) and in CSF (r=-0.686; p<0.001 for p-tau212; and r=-0.511; p<0.001 for p-tau181).

### Plasma p-tau212 levels are increased in asymptomatic Down syndrome

Plasma p-tau212 concentration was 2.4x times higher in the aDS group compared with cognitively normal (CN) euploid people (p<0.001) whereas plasma p-tau181 was not significantly changed (p=0.052).

### Plasma and CSF p-tau212 increase along the AD continuum in DSAD and sporadic AD

Both in individuals with DS and euploid participants, p-tau212 levels were increased in symptomatic patients in comparison with asymptomatic individuals. (Fig. 2A). For pDS we observed a 3.4x (p=0.003) mean fold increase in plasma and a 5.6x mean fold-fold increase in CSF in reference to aDS. For dDS we observed a 2.9x (p=0.004) mean fold increase in plasma and a 7.1x mean fold increase in CSF. For MCI-AD, compared with cognitively normal euploid people, we observed a 3.0x (p<0.001) mean fold increase in plasma and a 7.8x mean fold increase in CSF. AD dementia patients had a 3.8x mean fold increase in plasma (p<0.001) and a 9.1x mean fold increase in CSF. P-tau181 concentrations were also increased, but with a lower magnitude than p-tau212, and comparison between DS groups showed no significance (Fig. 2B). Both biomarkers kept the pattern of greater increases in CSF than in plasma (Supplementary Fig. 1A-B).

**Figure 2.**
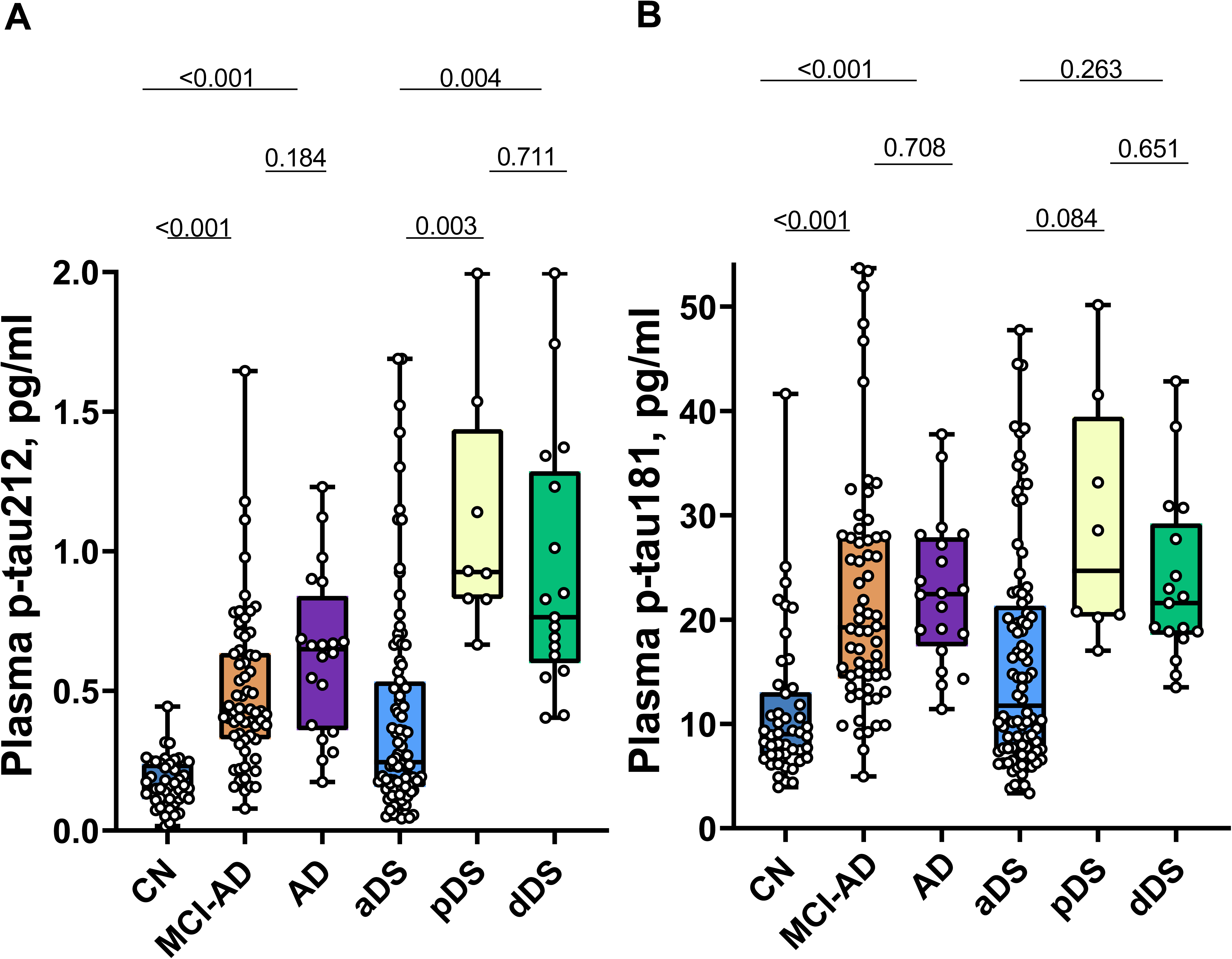
Plasma p-tau212 and p-tau181 levels in euploid and Down Syndrome (DS) groups. Box plots represent median and IQR, and boundaries of the whiskers are minimum to maximum values for **A)** plasma p-tau212 and **B)** plasma p-tau181. Differences for euploid participants are calculated for Mild Cognitively impaired Alzheimer’s Disease (MCI-AD; n=62) and Alzheimer’s Disease Dementia (AD; n=20) in reference to Cognitively Normal (CN; n=46) participants. Differences for prodromal Alzheimer’s Disease in DS (pDS; n=8) and Alzheimer’s Disease dementia in DS (dDS; n=17) are calculated in reference to asymptomatic (aDS; n=92). Age-adjusted analysis of covariance (ANCOVA) followed by Tukey post-hoc test is used to calculate differences across groups.

### Plasma p-tau212 has greater diagnostic accuracy than p-tau181

ROC analysis was used to evaluate the diagnostic performance of plasma and CSF p-tau212 and p-tau181. AUCs were usually higher for biomarkers in CSF, and for p-tau212 than for p-tau181. In our comparisons, we included Age+ Apolipoprotein E4 (APOE4)+Sex since they have been shown to influence diagnostic accuracy(30). AUCs of Age+APOE+Sex were not significantly different from p-tau212 but better than p-tau181. Both biomarkers had high accuracy to differentiate between CN and AD in plasma AUC = 0.96 (95% CI 0.92-1) for p-tau212 and AUC = 0.89 (95% CI 0.84-0.95) for p-tau181 (Fig. 3B). For differentiating between CN and MCI-AD (Fig. 3A) or MCI-AD+AD (Fig. 3C), p-tau212 had accuracy of 0.91 (95% CI 0.85-0.97) and AUC=0.93 [95% CI 0.88-0.97] respectively. That accuracy was significantly higher than p-tau181 - (p=0.026 for both comparisons). Plasma p-tau212 reached AUC=0.91 (95% CI 0.86-0.97) to differentiate between aDS and dDS diagnosis (Fig. 3D).

**Figure 3.**
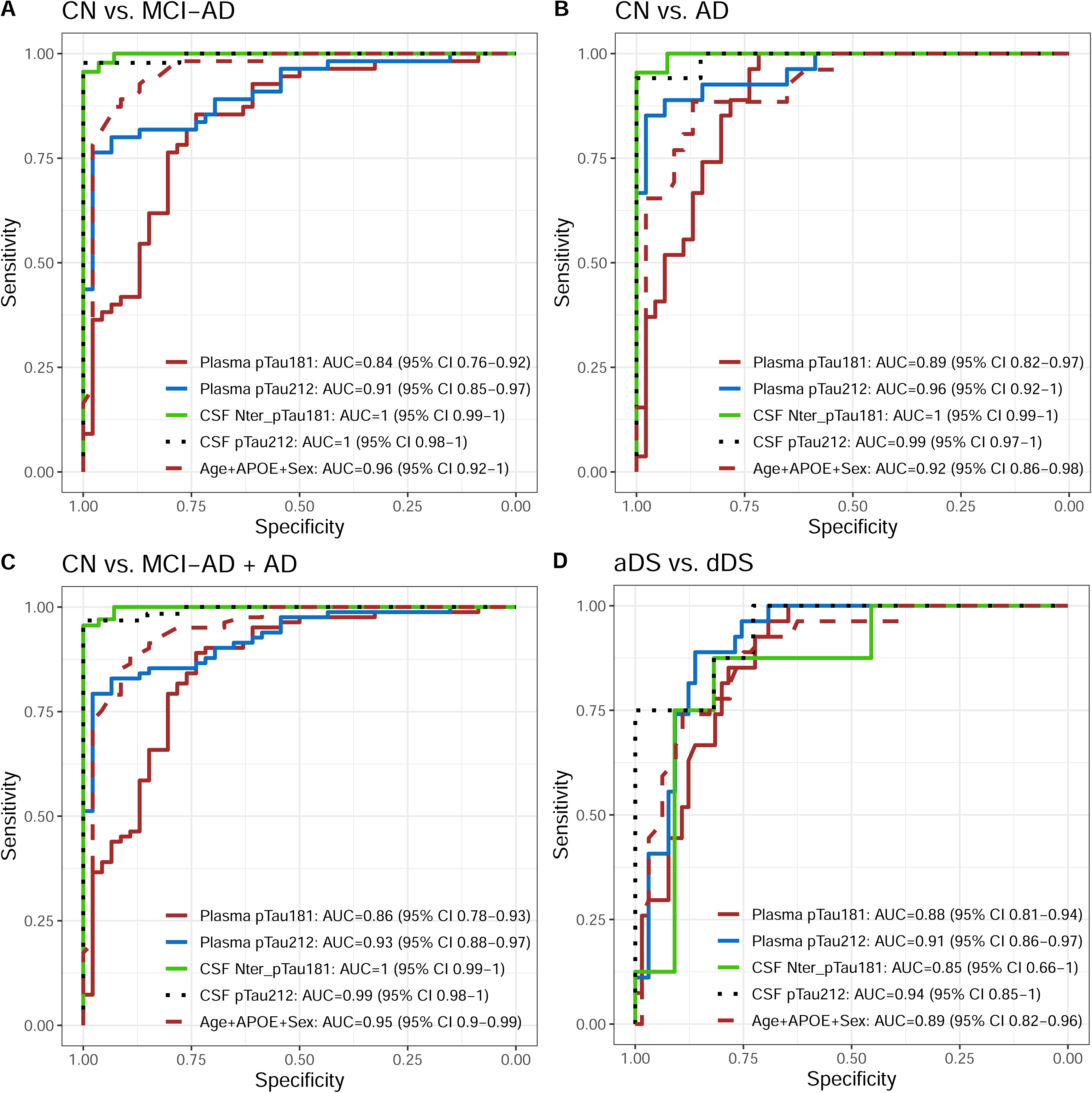
Diagnostic accuracy of plasma and CSF biomarkers to discriminate between sporadic and DSAD groups. P-tau212 and p-tau181 receiver operating characteristic curves (ROC) to discriminate between sporadic and DSAD groups. Plasma p-tau181, plasma p-tau212, CSF p-tau181, CSF p-tau212, and Age+APOE+Sex are on each graph. In **A**) ROC curves for differentiating Cognitively normal (CN) and Mild Cognitive Impairment - Alzheimer’s Dementia (MCI-AD). **B**) ROC curves to discriminate between CN and Alzheimer’s Disease (AD). **C**) ROC curves to differentiate between CN and MCI-AD combined with AD group. **D**) ROC curves to discriminate between asymptomatic Down syndrome (aDS) and prodromal Down Syndrome (pDS) + dementia Down syndrome (dDS).

### Plasma p-tau212 increases approximately parallel to amyloid PET positivity and has great accuracy in discriminating between Aβ+ and Aβ-participants

Both biomarkers had significant accuracy in discriminating between Aβ+ and Aβ-participants (Fig. 4). Plasma p-tau212 had numerically higher accuracy than p-tau181 both for DS (AUC = 0.86 (95%CI = 0.7-1) (Fig. 4B) and euploid participants AUC = 0.9 (0.83 – 0.96) (Fig. 4C).

**Figure 4.**
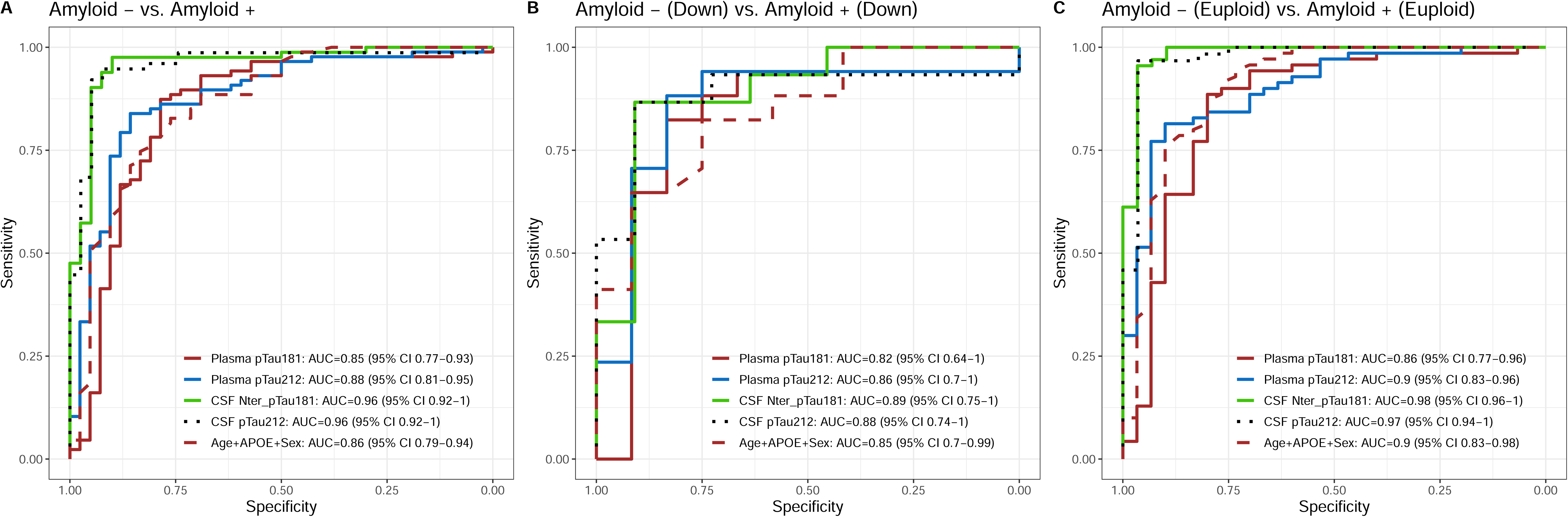
Diagnostic accuracy of plasma and CSF biomarkers to discriminate between Aβ+ and Aβ-participants. P-tau212 and p-tau181 receiver operating characteristic curves (ROC) to discriminate between amyloid positive (Aβ+) and amyloid negative (Aβ-) participants in sporadic and DSAD groups. Plasma p-tau181, plasma p-tau212, CSF p-tau181, CSF p-tau212, and Age+APOE+Sex are on each graph. In **A**) ROC curves for differentiating Aβ+ from Aβ-in whole cohort. **B**) ROC curves to discriminate Aβ+ and Aβ-in DS groups. **C**) ROC curves to differentiate between Aβ+ and Aβ-in euploid groups.

An early increase in plasma levels was observed many years before the onset of clinical AD symptoms in DS (Fig 5). For p-tau212 the increase started approximately when people became amyloid PET-positive, *i.e.*, in their late 30s, and approximately 15 years before the disease onset (Fig. 5A). P-tau181 started increasing approximately 10 years before the estimated disease onset (Fig. 5B).

**Figure 5.**
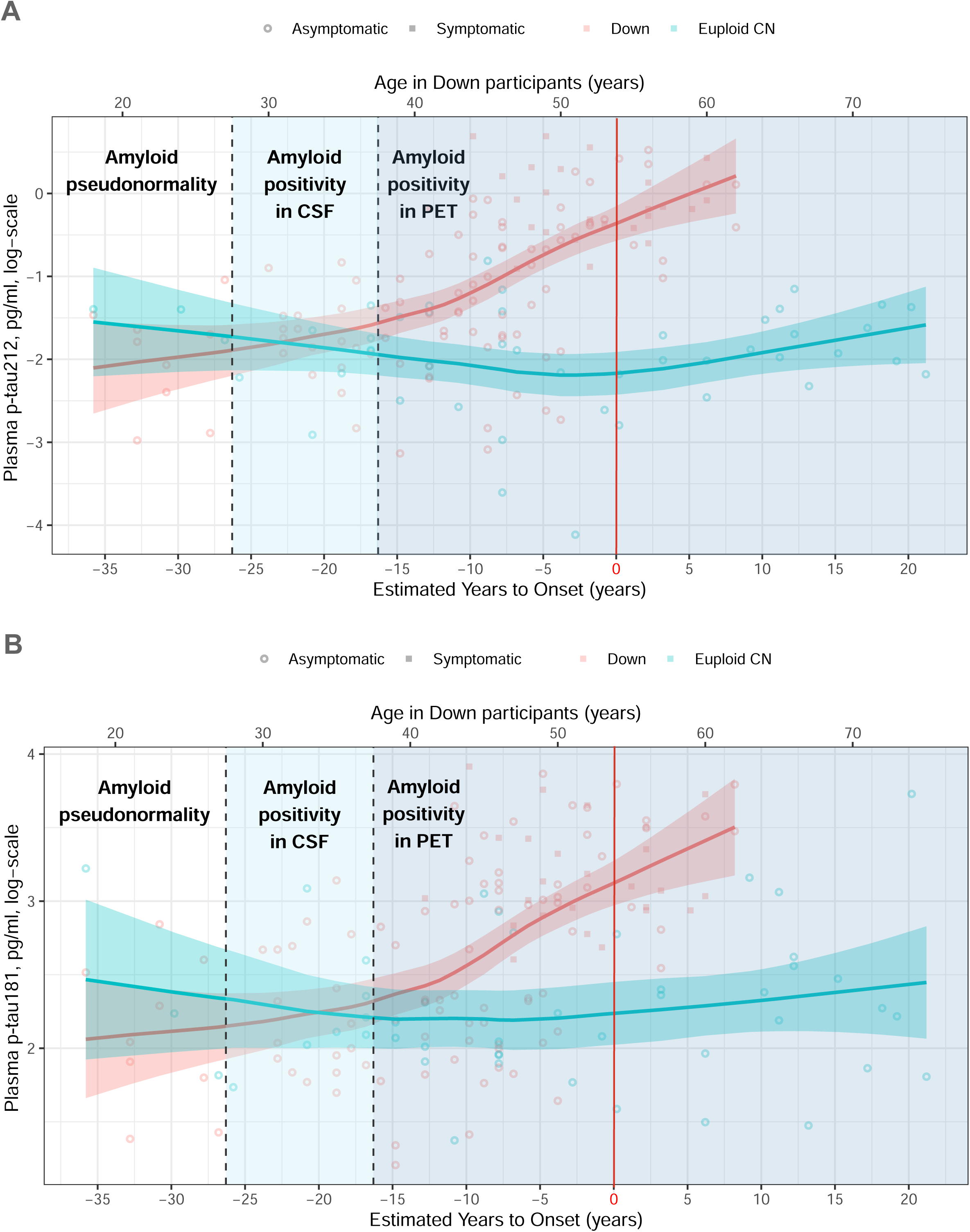
Age-related plasma p-tau212 and p-tau181 changes in Down syndrome and euploid controls. Open Circles represent asymptomatic and filled circles represent symptomatic participants. Down syndrome population is represented in red and cognitively normal euploid people are represented in blue. Horizontal **l**ines depict each group’s fitted loess model, and fainted bands display confidence intervals. The vertical red line represents the estimated years to symptom onset which is 53.8 for Down syndrome participants and was used as reference in euploid controls for comparison purposes.

## Discussion

In this cross-sectional study, we show that p-tau212 serves as a biomarker to track AD-related changes in sporadic cases and DSAD. P-tau212 has a very high correlation between plasma and CSF. The strongest correlation was observed for the DS population, where p-tau212 reached a greater correlation than for euploid participants. For p-tau181 correlation in euploid and DS groups was similar, suggesting greater translation from CSF to plasma for p-tau212 in DS populations.

We observed elevated p-tau212 concentration in plasma and CSF in aDS compared with euploid CN. That elevation is greater than the p-tau181 elevation, which did not reach statistical significance. Additionally, other reported biomarkers, such as p-tau217 Glial fibrillary acidic protein (GFAP)(30,31), had lower increases. This suggests the effects of DYRK1A gene dose interacting with AD pathology on p-tau212 levels in DS and a better fit for the use of this biomarker in the DS population.

Levels of plasma p-tau212 were significantly higher across disease groups. Plasma p-tau212 reached significance levels to differentiate between aDS vs pDS and aDS vs dDS groups while p-tau181 failed to do it. However, we think this might be a limitation of the small sample size used in this cohort since p-tau181 levels were previously shown to be significantly increased in pDS and aDS(30,32). In MCI-AD and AD dementia groups, we observed significant increases for both biomarkers, concomitantly having fold changes higher for p-tau212 which confirms our previous findings(22).

The excellent performance of the assay was confirmed in discriminating patients according to the diagnosis in both DS and euploid groups. P-tau212 additionally has greater accuracy than p-tau181 in discriminating between MCI-AD and control groups, simultaneously reaching 0.96 AUC to differentiate CN from AD. Additionally, plasma p-tau212 acquired very high AUC-ROC in discriminating between Aβ+ and Aβ-participants in both euploid and DS groups. This accuracy was not different from CSF accuracy, supporting the high between-matrix translation of p-tau212 and providing additional reasoning to use plasma p-tau212 to recruit participants for clinical trials.

Plasma p-tau212 starts increasing in the 30s, approximately when people start being positive in amyloid PET scans, and 5 years before we observe an increase in p-tau181. Additionally, the biomarker increased further as AD progressed towards symptomatic stages. Therefore p-tau212 could be useful to monitor the progression of asymptomatic DS people to prodromal AD. Moreover, the onset of the increase comes along with the appearance of neurofibrillary tangles (NFTs) and Aβ plaques in brain(4,33).

This is (to our best knowledge) the first study to measure p-tau212 in CSF and plasma as a biomarker for DSAD however, the potential involvement of p-tau212 in this population and its association with DYRK1A was published more than 20 years ago(13). The kinase is perceived as a target in DS and neurodegenerative diseases(8,34). Its under-or overexpression leads to different clinical phenotypes including cognitive impairment(34). Since DYRK1A is a dose-sensitive protein in which down-regulation or up-regulation has a critical role, DYRK1A inhibitors have already been widely explored in clinical trials and have been proven to improve cognitive function in DS people(10,35,36). Importantly, p-tau212 has already been successfully used to test the efficacy of DYRK1A inhibitor in cell models(37). The use of the chosen inhibitor was further shown to reverse the upregulation of p-tau212 in hippocampal tissue and temporal cortex in mouse models(37). Our novel plasma p-tau212 immunoassay provides a simple-to-implement and cost-effective opportunity to monitor the efficacy of DYRK1A inhibitors, or in the future – enhancers, not allowing the activity of this kinase to be reduced or increased to levels that could cause more harm than good. This utility will be explored in our future research.

The major strength of this study is the confirmation that p-tau212 is increased in the DS population, and levels of this biomarker increase with progression to AD dementia. P-tau212 reaches very high accuracy to differentiate between control and disease groups and Aβ+ and Aβ-participants. The high correlation between plasma and CSF p-tau212 also supports a very high translation of the results from CSF to plasma. DeLong tests between DS groups did not show any significantly better performance of CSF p-tau212 compared with plasma p-tau212 providing further evidence that plasma measurements can be used for clinical evaluation of AD pathophysiological processes occurring in patients with suspected disease. Advantages would also be reflected in the economy, availability, and perception of the test, since lumbar punctures or PET scans are costly, require resources, and might be perceived as frightening(20). Next, plasma p-tau212 increased approximately when people are starting to be amyloid PET positive and 5 years before p-tau181, indicating the benefits in disease monitoring.

This study has a few limitations. First is a slightly low representation of prodromal-DS participants, which prohibits us from making better AUC-ROC analysis in that group. Ideally, longitudinal measurements of p-tau212 in the DS population would tell us more about the trajectories of this biomarker. The second limitation is that p-tau217 measurements are unavailable at the moment, however direct comparison between biomarkers was not a purpose of experiments presented in this article. Still, p-tau181 is the most commercialized and fully automated immunoassay, with great utility in AD.

## Conclusions

In conclusion, we have confirmed that levels of plasma p-tau212 are increased in the DS population and sporadic AD cases including prodromal and MCI states. High accuracy in discriminating amyloid positive from amyloid negative people and increase in parallel to amyloid-PET positivity give the promise to evaluate ongoing pathophysiological AD processes many years before the disease onset in individuals with DS. This will also facilitate participants recrutation for clinical trials. This is a cost-effective application that provides higher chance to receive appropriate therapy. Plasma p-tau212 will also find high utility for theragnostic, to monitor therapy efficacy, and as a target engagement biomarker in clinical trials both in sporadic and DSAD.

## Data Availability

Blinded Anonymized data is available on reasonable request from the corresponding author. Request will be reviewed by the investigators and respective institutions to verify if data transfer is in the agreement with EU legislation on the general data protection or is subject to any intellectual property or confidentiality obligations.

## Abbreviations

AD: Alzheimeŕs disease
DS: Down Syndrome
DYRK1A: dual-specificity tyrosine phosphorylation-regulated kinase 1A
p-tauX: tau phosphorylated at amino acid X
DSAD: Down Syndrome Alzheimeŕs disease
CSF: Cerebrospinal Fluid
AUC-ROC: Area Under the Curve and Receiver Operating Curves
ANCOVA: Age-adjusted analysis of covariance
LOESS: locally estimated scatterplot smoothing
pDS: prodromal Down syndrome
aDS: asymptomatic Down syndrome
dDS: dementia Down syndrome
PET: Positron Emission Tomography
MCI: mild cognitive impairment
Aβ: amyloid beta
APP: amyloid precursor protein
GSK-3β: Glycogen synthase kinase-3β
PP1A: Protein phosphatase 1A
DABNI: Down Alzheimer Barcelona Neuroimaging Initiative
SPIN: Sant Pau Initiative on Neurodegeneration
CN: cognitively normal
IQR: interquartile range
SD: standard deviation
CAMCOG-DS: The Cambridge Cognitive Examination adapted for individuals with Down Syndrome
APOE4: Apolipoprotein E4
GFAP: Glial fibrillary acidic protein
NFTs: Neurofibrillary tangles

## Declarations

### Ethics approval and consent to participate

Study procedures were approved by the Sant Pau Ethics Committee (IIBSP-NGF-2018-36 and IIBSP-DOW-2014-30), following the standards for medical research in humans, as recommended in the Declaration of Helsinki. All participants or their legally authorized representative gave written informed consent before enrolment.

### Consent for publication

Not applicable

### Authors’ contributions

PRK, DA, NJA, HZ, TKK, JF and KB created the concept and design. Data acquisition and analysis was performed by PRK, DA, LMG, JLR, NJA, JF. SF, LM, BB, MT, IB, PH, LV, AL, MC-I contributed to the sample selection/and or interpretation of the data. PKK, DA, LMG, JLR, FG-O, HZ, TKK, JF and KB drafted the manuscript, and all authors revised. All authors read and approved the final manuscript.

## Acknowledgements

We thank all the participants with Down syndrome, their families, and their carers for their support of, and dedication to this research. We also acknowledge Fundació Catalana Síndrome de Down for global support and the members of the Alzheimer Down Unit and the Memory Unit from Hospital de la Santa Creu i Sant Pau for their daily work and dedication.

