## Supplementary file for "Plasma p-tau212 as a biomarker of sporadic and Down Syndrome Alzheimer’s disease"

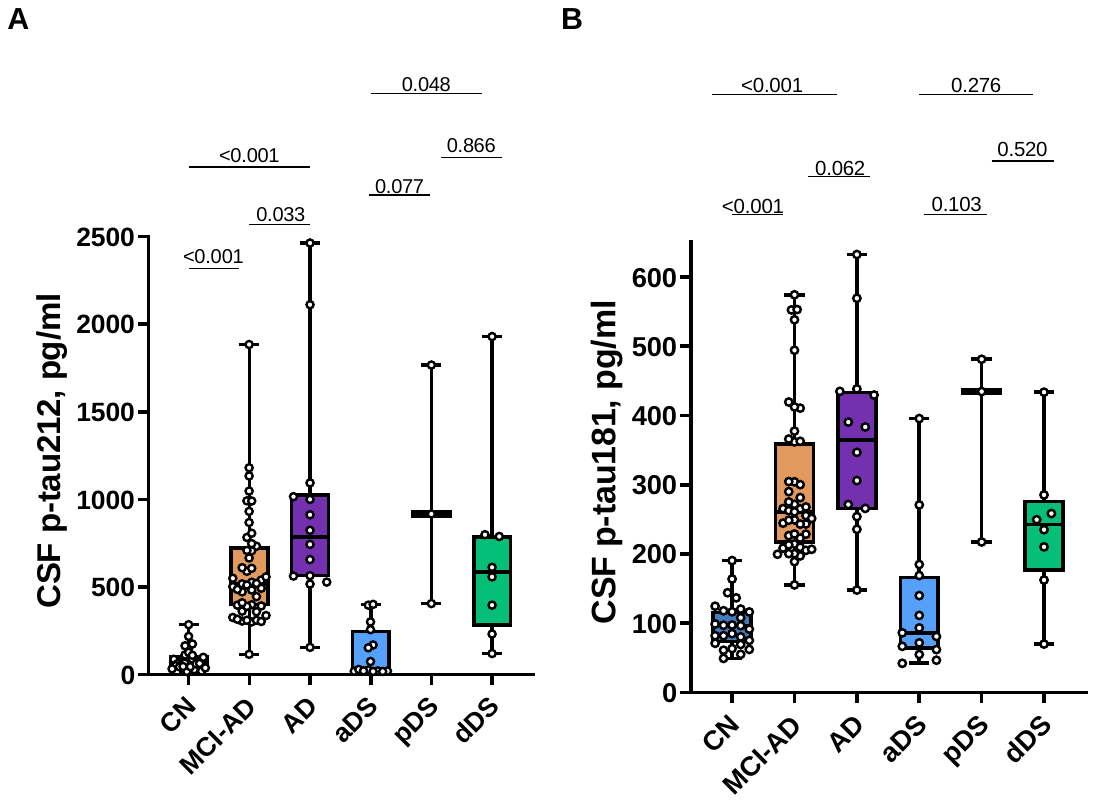

**Supplemenatary Fig. 1 CSF p-tau212 and p-tau181 levels in euploid and Down Syndrome (DS) groups.** Box plots represent median and IQR, boundaries of the wishkers are minimum to maximum values for **A)** CSF p-tau212 and **B)** CSF p-tau181. Differences for euploid participants are calculated for Mild Cognitively impaired Alzheimer’s Disease (MCI-AD; n=47) and Alzheimer’s Disease Dementia (AD; n=14) in reference to Cognitively Normal (CN; n=27) participants. Differences for prodromal Alzheimer’s Disease in DS (pDS; n=3) and Alzheimer’s Disease dementia in DS (dDS; n=8) are calculated in reference to asymptomatic (aDS; n=15). Age adjusted analysis of covariance (ANCOVA) followed by Tukey post-hoc test is used to calculate differences across groups.

**Supplemenatry Table 1. CSF Biomarker levels**

|  | aDS | pDS | dDS | CN | MCI-AD | AD |
| --- | --- | --- | --- | --- | --- | --- |
| N= | 15 | 3 | 8 | 27 | 47 | 14 |
| CSF p-tau212 range | 19.3-402 | 406-1768 | 121-1930 | 18.9-286 | 116 - 1885 | 156 - 2464 |
| CSF p-tau212 mean (SD) | 129 (144) | 1031 (688) | 680 (561) | 88.44 (64.2) | 594.9 (310) | 940 (627) |
| CSF p-tau212 median[IQR] | 30.5 [21.5-257] | 918 [406-1768] | 586 [274-797] | 64.4 [44.7-113] | 518 [390-733] | 784 [555-1036] |
| CSF p-tau181 range | 42-395 | 217-481 | 70-434 | 49.2 - 192 | 155 - 362 | 148 – 436 |
| CSF p-tau181 mean (SD) | 125 (97.5) | 378 (141) | 238 (104) | 98.4 (34) | 294 (107) | 365 (132) |
| CSF p-tau181 median[IQR] | 86.1 [61.9-169] | 435 [217-481] | 242 [174-279] | 96.5 [71.0-118] | 261 [214-362] | 365 [263-436] |

aDS – asymptomatic Down syndrome; pDS – prodromal Down syndrome; dDS – dementia Down syndrome; CN – cognitively normal; MCI-AD – mild cognitive impairment due to Alzheimer’s Disease; AD – Alzheimer’s disease dementia; IQR – interquartile range; SD – standard deviation.
